# Timely referral for advanced therapy in Parkinson’s disease. Development of a screening tool

**DOI:** 10.1101/2021.02.10.21251496

**Authors:** Harmen R. Moes, Jolien ten Kate, Axel T. Portman, Barbera van Harten, Mirjam E. van Kesteren, Tjeerd Mondria, Gerton Lunter, Erik Buskens, Teus van Laar

## Abstract

**Objectives:** To develop a screening tool for timely referral for advanced therapy (AT) in patients with Parkinson’s disease (PD), and to compare the newly-developed tool with the published 5-2-1 criteria.

**Design:** Cross-sectional, diagnostic, observational study and multivariable logistic regression analysis for item selection.

**Setting:** 8 hospitals in the Northern part of the Netherlands situated in the catchment area of a specialized movement disorder centre.

**Participants:** 259 consecutive PD patients not yet on AT visiting the outpatient clinic of participating hospitals from February 2017 to July 2018.

**Predictors:** 24 patient and disease characteristics as assessed by the treating neurologists, and scores on the NMS questionnaire.

**Outcome:** Apparent eligibility for referral for AT based on consensus by a panel of 5 experts in the field of AT.

**Results:** 17 patients were deemed eligible for referral for AT (point prevalence: 6.6%). Presence of response fluctuations and troublesome dyskinesias were the strongest independent predictors of being eligible for referral. Both variables were included in the final model, as well as levodopa equivalent daily dose. Decision curve analysis showed that the new model outperformed the 5-2-1 criteria. A simple chart was constructed to provide guidance for referral. Discrimination of this simplified scoring system was good (AUC after bootstrapping: 0.97).

**Conclusion:** The screening tool may improve efficiency of referral and subsequent treatment with AT in patients with PD. External validation is required prior to application in daily practice.

Anyone who wishes to share, reuse, remix, or adapt this material must obtain permission from the corresponding author.

## INTRODUCTION

Parkinson’s disease (PD) is a common neurodegenerative disorder for which a cure is lacking.[1] Treatment is aimed at alleviating symptoms and improving quality of life through lessening of motor and non-motor symptoms using dopaminomimetic drugs.[2] Within ten years of treatment with oral or transdermal dopaminomimetics, many PD patients develop disabling motor complications, typically involving (unpredictable) response fluctuations and troublesome dyskinesias.[3] Initially, these motor complications can be circumvented by optimizing the treatment regimen, but at some point oral and transdermal treatment no longer suffices. [4]

Advanced therapies (AT) are effective treatments in patients with PD suffering from motor complications that can no longer be treated adequately using oral and transdermal dopaminergic drugs.[5] The term AT refers to three invasive treatment modalities, namely deep brain stimulation of the subthalamic nucleus (DBS), continuous subcutaneous apomorphine infusion (CSAI) and infusion of levodopa-carbidopa intestinal gel (LCIG).[6–9] All three treatments have been shown to improve motor function and increase quality of life in PD patients suffering from unpredictable response fluctuations and troublesome dyskinesias.[7–9]

Patient eligibility for AT is usually determined in specialized movement disorders centres by means of a comprehensive and standardized selection process. [10,11] Currently, however, timely referral to these specialized centres is suboptimal, as it remains challenging to identify PD patients who might be candidates for AT. Indeed, neurologists who have less experience with AT and infrequently treat patients with PD, have difficulty timing referral, i.e., not too early and not too late.[Moes *et al*, in submission]. This difficulty is probably due to a lack of clear and easy-to-use criteria for pre-selection.[12] This leads to both inappropriate referrals (i.e. a patient will be rejected in the specialized centre) and under-referral (i.e. a potentially eligible patient is not given the opportunity to be evaluated).[13] With regard to the suboptimal referral pattern, a group of international experts pleaded for the development of guidance for general neurologists to improve timely referral of patients for AT.[14]

Recently, multiple attempts have been made to define criteria for which patients should be referred for AT.[11,15] These attempts have led to the development of a simple screening tool, the so-called 5-2-1 screening criteria, to identify advanced PD patients in whom any AT may be considered.[14,16,17] According to these criteria patients who fulfil at least one of the following characteristics have advanced PD: 5 times oral levodopa tablets taken daily; 2 hours of off time per day; 1 hour of troublesome dyskinesia per day. Although these criteria are easy to use and possibly sensitive in diagnosing advanced PD,[16,17] the 5-2-1 criteria have not been developed nor tested using prospective data from routine care. An important limitation is the lack of data on the diagnostic accuracy of the 5-2-1 criteria, including a reliable estimation of the positive predictive value.

To circumvent the limitations of the 5-2-1 criteria, the aim of this study was to develop a new, user-friendly screening tool using data from a cross-sectional study in routine care. An additional goal was to compare the diagnostic performance of the newly developed tool with the 5-2-1 criteria.

## METHODS

The TRIPOD guidelines for multivariable prediction models were followed for the development, internal validation and reporting of this clinical screening tool. [18] The Medical Ethics Review Committee of the University Medical Center Groningen (the Netherlands) waived the need for consent. All patient data was processed in accordance with current privacy legislation.

### Design

This was a cross-sectional study aimed at the development of a tool to screen for PD patients eligible for referral for AT. The target population were consecutive PD patients attending regular outpatient appointments to secondary care settings. The study was conducted between February 2017 and November 2018. The clinical data was collected prospectively in the period February 2017-July 2018. The assessments by the expert panel took place in three meetings between June 2018 and November 2018. No external validation was performed.

### Participants

The study was be performed in a pre-existing referral network in the northern part of the Netherlands (Parkinson Platform Noord Nederland). This is a network care setting with one university hospital (University Medical Center Groningen, offering DBS, CSAI and LCIG) and 13 referring community hospitals.

In short, characteristics of consecutive PD patients visiting the outpatient clinic were assessed and recorded by neurologists and supervised PD nurses (health care professionals; HCP) from the community hospitals. Hospitals participating in the study were instructed to include the first 50 consecutive PD patients. A panel of experts evaluated the anonymized patient data to determine whether individual patients would be good candidates for referral (reference test).

### Patients and HCP

Inclusion criteria for PD patients were age > 18 years and levodopa-responsive PD according to the MDS Diagnostic Criteria.[19] Patients who had already been treated with AT were excluded. In the community hospitals, HCP were entitled to participate in the data collection if they were officially registered as a general neurologist, or as PD nurse provided that he/she was supervised by a neurologist. Neurologists who were member of the expert panel were not allowed to be involved in the initial assessment of the patients enrolled.

### Expert panel

The expert panel consisted of five movement disorder specialists (neurologists) with extensive experience in treating PD patients and substantial experience in applying AT. Table A1 (appendix) shows the characteristics of the experts. One neurologist (TvL) was expert in the field of DBS, LCIG and CSAI. Two neurologists (MvK and BvH) were experts in the field of LCIG infusion. Two other neurologists (AP and TM) were experts in the field of CSAI. The experts did not receive any financial compensation for participating in this study.

### Outcome and predictor variables

The screening tool was designed to predict the following outcome: eligibility for referral to a specialized centre for additional evaluation for treatment with any AT (DBS, LCIG or CSAI). In short, the outcome was determined by an expert panel (see below). The potential predictors of the model were clinical and demographic variables as recorded during routine consultations at the outpatient clinic.

### Predictor variables

All included patients were asked to complete the Dutch version of the NMS-quest prior to their regular visit to the outpatient neurology clinic.[20] During the visit, the HCP assessed the clinical and demographic variables of interest and filled out the study assessment form (see table A2 in the appendix for the complete list of all potential predictors). The form contained information on demographics, motor symptoms, non-motor symptoms and the medication regimen, all considered relevant in determining eligibility for AT.[14] The assessment of these variables reflected daily practice, as all information of interest was easily accessible during routine consultations at the outpatient clinic.

In addition to filling out the clinical characteristics, the HCP had to state whether they felt the patient would be eligible for referral for AT.

### Outcome

In the absence of a standardized reference test for eligibility for referral (the ‘gold standard’), the primary outcome was based on the judgment (consensus) of an expert panel. For practical reasons, we used a staged approach for decision making, as described elsewhere. [21]

The expert panel evaluated all the recorded information on the enrolled patients (see section predictor variables), while they remained blinded for the judgment of the HCP. Based on all this information, the expert panel determined whether the patient was eligible for referral for AT.

First, all experts assessed each case individually and submitted their judgement to the main investigator (HM). Second, a selection of cases was discussed during a plenary session with the experts. Cases were selected if there was disagreement among the individual experts, or disagreement between the judgment of the HCP and the expert panel (see figure A1 in appendix). During the plenary session, consensus on the selected cases was reached using voting rounds. All experts voted by raising a ballot paper (yes, no, indifferent). In case of no unanimity, the experts discussed their viewpoints. Thereafter, a second voting round was performed. In case of no consensus, a majority vote was decisive.

### Sample size calculation

A common maxim for group size calculations in studies developing prediction models is that one requires at least 10 cases (from the smallest category) per predictor variable.[18,22] Our initial aim was to obtain a group size of 1000 patients in total. However, for practical reasons, we downsized this aim to 300 patients. We assumed this number to be sufficient to study 3 independent predictors, as we estimated the prevalence of AT eligibility to be 10% among all PD patients.

### Data processing

The data collection and management for this research project was performed using the OpenClinica, Community Edition, version 3.14. All study data was stored in a secure location to which only authorized researchers had access. The data set from OpenClinica was exported to a csv-file to perform the statistical analyses.

### Statistical analyses

The statistical analyses were performed in SPSS Statistics version 25.0, R Statistics version 4.0.2 and Microsoft Excel 2019.

Patient demographics were examined using descriptive statistics. Continuous, normally distributed variables were summarized as mean with standard deviation (SD). Categorical and ordinal variables were described as number and percentage.

The prevalence of referral eligibility for AT was determined using the Clopper-Pearson binomial test at a 95% confidence interval (95% CI). The degree of agreement between the assessing HCP and the expert panel was expressed with Cohen’s kappa.

### Missing values

Missing values were not imputed in the primary, univariable logistic regression analyses. Prior to multivariable logistic regression (see modelling), missing values were imputed using multiple imputation (MICE package in R). In total, 10 imputation sets were created. Missing values were predicted on the basis of all other predictors that had at least a minimum correlation (0.2) to make sure the imputation model would converge. The outcome was excluded from the imputation model. For continuous variables, we checked that observed and imputed values were in the same range.

### Modelling, predictor selection and internal validation

Associations between potential predictors and the outcome variable were studied using univariable logistic regression analyses. Continuous variables were not dichotomized. Ordinal variables were not converted into continuous variables. P-values < 0.05 were considered statistically significant. No adjustments were made for multiple testing.

Before the multiple regression modelling, small categories of ordinal predictors were grouped to eliminate sparse categories. During multiple logistic regression modelling, key predictors were identified using 50-fold cross validation and lasso GLM in order to internally validate the model and minimize the risk of overfitting. We used a conservative estimate for lambda (1 SE) to find the simplest model by means of ensemble modelling and majority voting.

### Model performance, simplified scoring system and comparison with the 5-2-1 criteria

We reported the β values and odds ratios of the final multivariable logistic regression model (‘pooled’ estimate of the multiple imputed dataset). For the variable levodopa equivalent daily dose (LEDD)[23], the β values and odds ratios were reported with increments of 100 mg/day. To create an easy-to-use screening tool, we made a simplified version of the model by rounding the regression coefficients to easy-to-sum integers.

A calibration plot was constructed to evaluate the calibration of the final model. We assessed the predictive performance of the simplified screening tool by examining the receiver operating characteristics (ROC) curve and its area under the curve (AUC) in the multiple imputed dataset. We reported a bootstrapped estimate of the AUC of the simplified scoring system. For bootstrapping, 1000 samples were drawn with replacement of the original sample.

We used decision curve analysis (DCA) compare the clinical utility of the final model with 5-2-1 criteria.[24] In short, DCA compares the net benefit of two models for a range of threshold values. At any given threshold, the model with the higher net benefit is the preferred model. To calculate the numbers for the 5-2-1 criteria, we used the imputed dataset and we determined that «at least 1 hour of troublesome dyskinesias» was positive if the patient had 0-2 hours of troublesome dyskinesias per day (on the study assessment form). Additional information on DCA, including a step-by-step guide for interpreting decision curves, can be found at www.decisioncurveanalysis.org.[25,26]

### Screening tool

For user-friendliness we created a graphical presentation of the simplified scoring system. To illustrate the possible application of the prototype screening tool, we obtained an arbitrary, non-empirical threshold value by determining the coordinates of the ROC-curve at which the sum of sensitivity and specificity was highest. In addition, we calculated the positive predictive value and negative predictive value.

## RESULTS

### Included cases

In total, 263 assessment forms and NMS questionnaires were filled out in eight different hospitals (figure 1). Of these, 259 cases were suitable for assessment by the expert panel.

**Figure 1.**
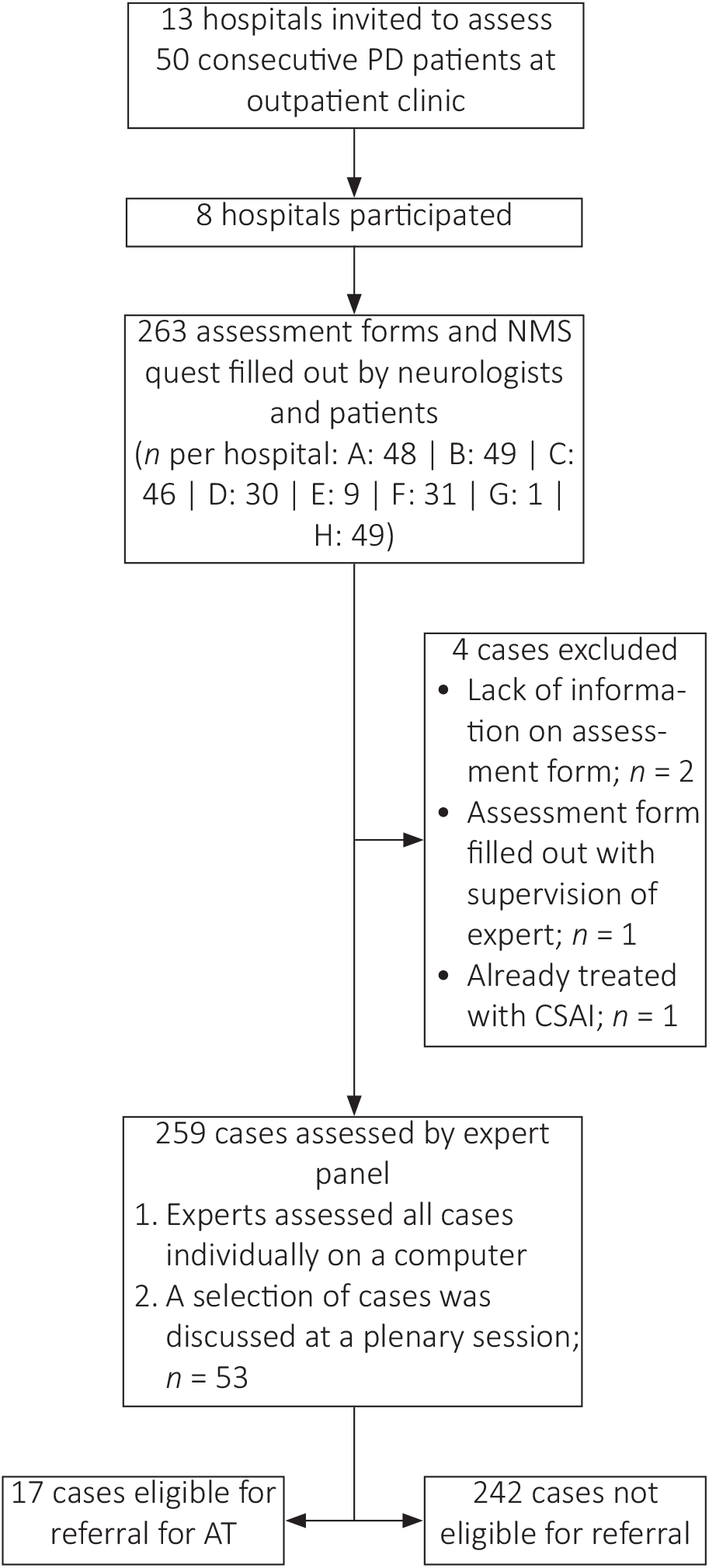
Patient flow chart. AT = advanced therapy; CSAI = continuous subcutaneous apomorphine infusion; NMS = non-motor symptoms; PD = Parkinson’s disease.

The HCP deemed 24 patients eligible for referral for AT, while 12 of these patients did not want to be referred for AT (figure A2 in appendix). The expert panel considered 17 cases to be eligible for referral for AT; the remaining 242 patients were not considered eligible for referral (figure A3). Based on these numbers, the prevalence of referral eligibility for AT was 6.6% (95% CI: 3.9-10.3%). The agreement between the assessment of the referring HCP and the expert panel was moderate (Cohen’s Kappa: 0.44, table A3 in appendix).

### Patient characteristics

The patient and disease characteristics of the included cases are shown in table 1. The mean age of all patients was 68.0 years (SD: 9.6). The majority of the patients were male (65.9%). The mean disease duration was 5.0 years (SD: 4.5), with the disease duration being significantly higher in the group of patients eligible for referral for AT (mean duration of disease: 11.5 years; SD: 3.1).

**Table 1.**
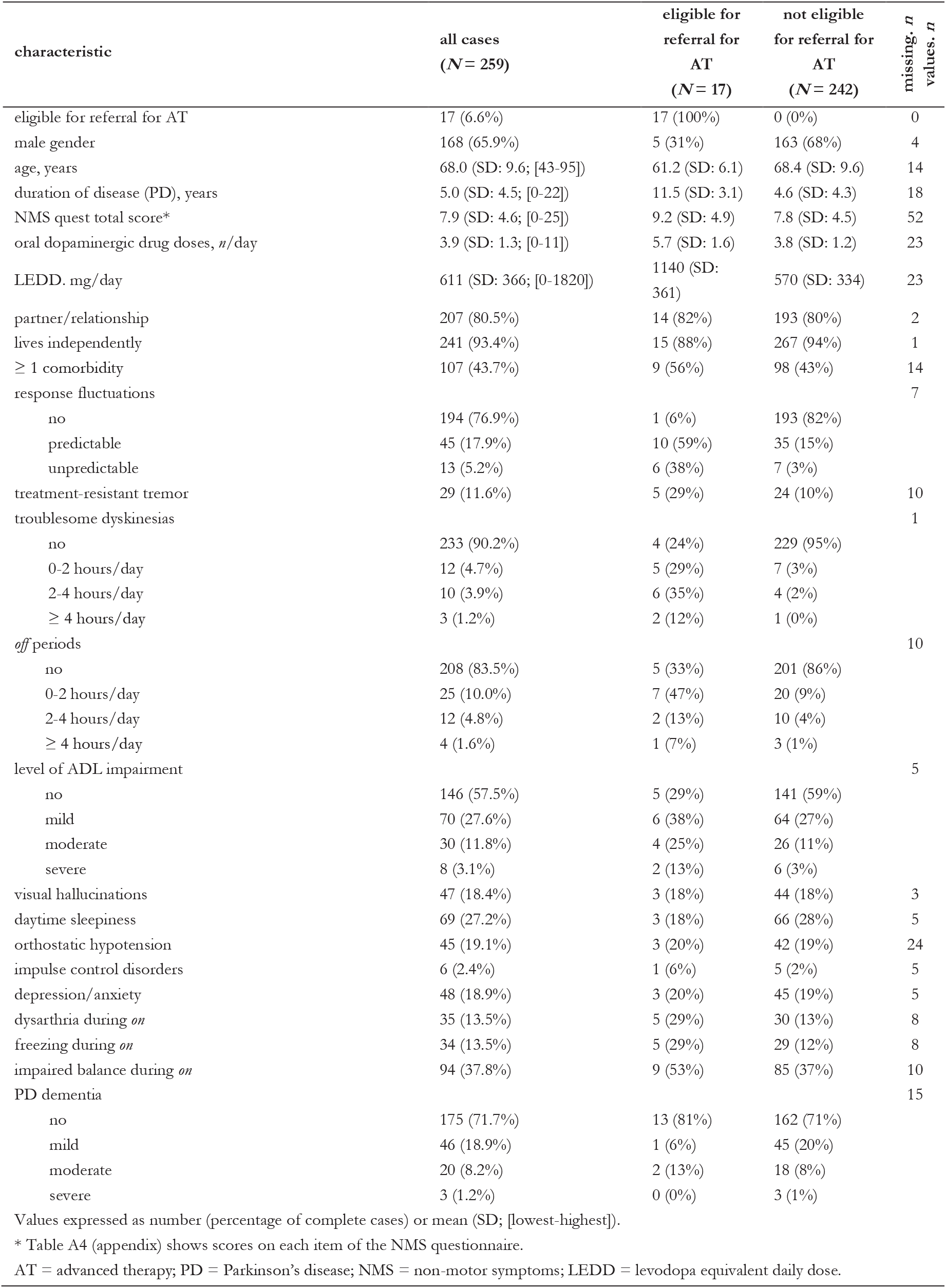
Patient and disease characteristics of the included cases

Motor symptoms of advanced PD were more common in patients eligible for referral for AT. However, these motor symptoms were absent in the majority of all patients: 76.9% had no response fluctuations, 88.4% experienced no treatment-resistant tremor, 90.2% did not suffer from troublesome dyskinesias, and 83.5% did not experience off periods.

### Model development and model specification

Table 2 shows the uncorrected, univariable association between each candidate predictor and eligibility for referral for AT. The number of complete cases and missing values per candidate predictor are also shown. Table 3 presents the final multivariable logistic regression model for eligibility for referral for AT, including the variables LEDD, response fluctuations and troublesome. Table 4 shows the simplified scoring system for determining eligibility for referral for AT.

**Table 2.** Univariable logistic regression analysis of the candidate predictors

| characteristic | odds ratio<br>(95% CI) | p-value* | complete/missing |  |
| --- | --- | --- | --- | --- |
|  |  |  | eligible<br>(N = 17) | not eligible<br>(N = 242) |
| male gender | 0.21 (0.07-0.63) | **0.005** | 16/1 | 239/3 |
| age, years | 0.92 (0.87-0.98) | **0.005** | 16/1 | 229/13 |
| duration of disease (PD), years | 1.29 (1.16-1.43) | **<0.001** | 16/1 | 225/17 |
| NMS quest total score** | 1.06 (0.95-1.20) | 0.310 | 12/5 | 195/47 |
| oral dopaminergic drug doses, n/day | 2.96 (1.86-4.70) | **<0.001** | 17/0 | 219/23 |
| LEDD, mg/day | 1.003 (1.002-1.005) | **<0.001** | 17/0 | 219/23 |
| partner/relationship | 1.14 (0.31-4.12) | 0.846 | 17/0 | 240/2 |
| lives independently | 0.50 (0.10-2.38) | 0.382 | 17/0 | 241/1 |
| ≥ 1 comorbidity | 1.72 (0.62-4.78) | 0.299 | 16/1 | 229/12 |
| response fluctuations |  |  | 17/0 | 235/7 |
| predictable | 55.1 (6.8-444.5) | **<0.001** |  |  |
| unpredictable | 165.4 (17.5-1565.3) | **<0.001** |  |  |
| therapy resistant tremor | 3.61 (1.17-11.13) | **0.025** | 17/0 | 232/10 |
| troublesome dyskinesias |  |  | 17/0 | 241/1 |
| 0-2 hours/day | 40.9 (9.0-186.0) | **<0.001** |  |  |
| 2-4 hours/day | 85.9 (17.2-427.7) | **<0.001** |  |  |
| ≥ 4 hours/day | 114.5 (8.5-1535.5) | **<0.001** |  |  |
| off periods |  |  | 15/2 | 234/8 |
| 0-2 hours/day | 14.1 (4.1-48.4) | **<0.001** |  |  |
| 2-4 hours/day | 8.0 (1.4-46.7) | **0.020** |  |  |
| ≥ 4 hours/day | 13.4 (1.2-152.3) | **0.036** |  |  |
| level of ADL impairment |  |  | 17/0 | 237/5 |
| mild | 2.64 (0.78-8.98) | **0.119** |  |  |
| moderate | 4.31 (1.09-17.24) | **0.037** |  |  |
| severe | 9.40 (1.51-58.72) | **0.017** |  |  |
| visual hallucinations | 0.95 (0.26-3.45) | 0.937 | 17/0 | 239/3 |
| daytime sleepiness | 0.56 (0.16-2.00) | 0.367 | 17/0 | 237/5 |
| orthostatic hypotension | 1.06 (0.29-3.92) | 0.931 | 15/2 | 220/22 |
| impulse control disorders | 2.90 (0.32-26.33) | 0.344 | 17/0 | 237/5 |
| depression/anxiety | 1.08 (0.29-3.98) | 0.911 | 15/2 | 239/3 |
| dysarthria during on | 2.83 (0.93-8.61) | **0.066** | 17/0 | 234/8 |
| freezing during on | 2.95 (0.97-8.97) | **0.057** | 17/0 | 234/8 |
| impaired balance during on | 1.95 (0.72-5.23) | **0.187** | 17/0 | 232/10 |
| PD dementia |  |  | 16/1 | 228/14 |
| mild | 0.27 (0.04-2.17) | 0.222 |  |  |
| moderate | 1.39 (0.29-6.63) | 0.684 |  |  |
| severe | 0.00 (0.00) | 1.00 |  |  |

\* P-values smaller than $p < 0.05$ are marked in red.

**Table 3.** Full model for testing eligibility for referral for an advanced therapy, including the intercept

| characteristic | B (SE) | odds ratio (95% CI) | p-value |
| --- | --- | --- | --- |
| intercept | -6.05 (1.30) |  |  |
| LEDD (100 mg/day) | 0.28 (0.10) | 1.32 (1.08-1.60) | 0.007 |
| response fluctuations (yes vs no) | 2.46 (1.15) | 11.7 (1.2-113.4) | 0.034 |
| troublesome dyskinesias (yes vs no) | 2.11 (0.57) | 8.24 (2.7-25.5) | < 0.001 |
| ROC area (95% CI) | 0.97 (0.94-0.99) |  |  |

SE = standard error; 95% CI = 95% confidence interval; LEDD = levodopa equivalent daily dose.

**Table 4.** Graphical representation of the simplified scoring system (screening tool) of the model as presented in table 3

|  | troublesome dyskinesias |  |  |  |
| --- | --- | --- | --- | --- |
|  | no |  | yes |  |
|  | no | yes | no | yes |
| 0 | 0.0 | 2.0 | 2.0 | 4.0 |
| 300 | 0.9 | 2.9 | 2.9 | 4.9 |
| 600 | 1.8 | 3.8 | 3.8 | 5.8 |
| 900 | 2.7 | 4.7 | 4.7 | 6.7 |
| 1200 | 3.6 | 5.6 | 5.6 | 7.6 |
| 1500 | 4.5 | 6.5 | 6.5 | 8.5 |
| 1800 | 5.4 | 7.4 | 7.4 | 9.4 |
| 2100 | 6.3 | 8.3 | 8.3 | 10.3 |
|  | no | yes | no | yes |
|  | response fluctuations |  |  |  |

A simplified scoring system to decide on referral of patients is: $0.3 \times \text{LEDD}^* + 2 \times \text{response fluctuations} + 2 \times \text{troublesome dyskinesias}$ .
\*Please note: the coefficient for LEDD was calculated with increments of 100 mg/day.
Numbers in the table represent calculated scores. Green cells have scores higher than the arbitrarily chosen cut-off point of 6.42. Green cells refer to patients who are eligible for referral for AT, according to the model.

### Performance of the model and screening tool

The calibration slope of the model (figure 2) showed moderate calibration in-the-large, with a tendency to overestimation. Decision curve analysis (figure 3) showed that the model had higher net benefit than the 5-2-1-criteria for all possible threshold values. The 5-2-1 criteria had a comparable net benefit for lower threshold values, i.e. when the ‘costs’ of false-negative are approximately 50 times higher than the ‘costs’ of a false-positive (threshold value: 2% [1/0.02 = 50])). The simplified scoring system (screening tool) had a good discriminative performance (ROC area: 0.97; 95% CI: 0.94-0.99; bootstrap bias: 0.0003; figure 4).

**Figure 2.**
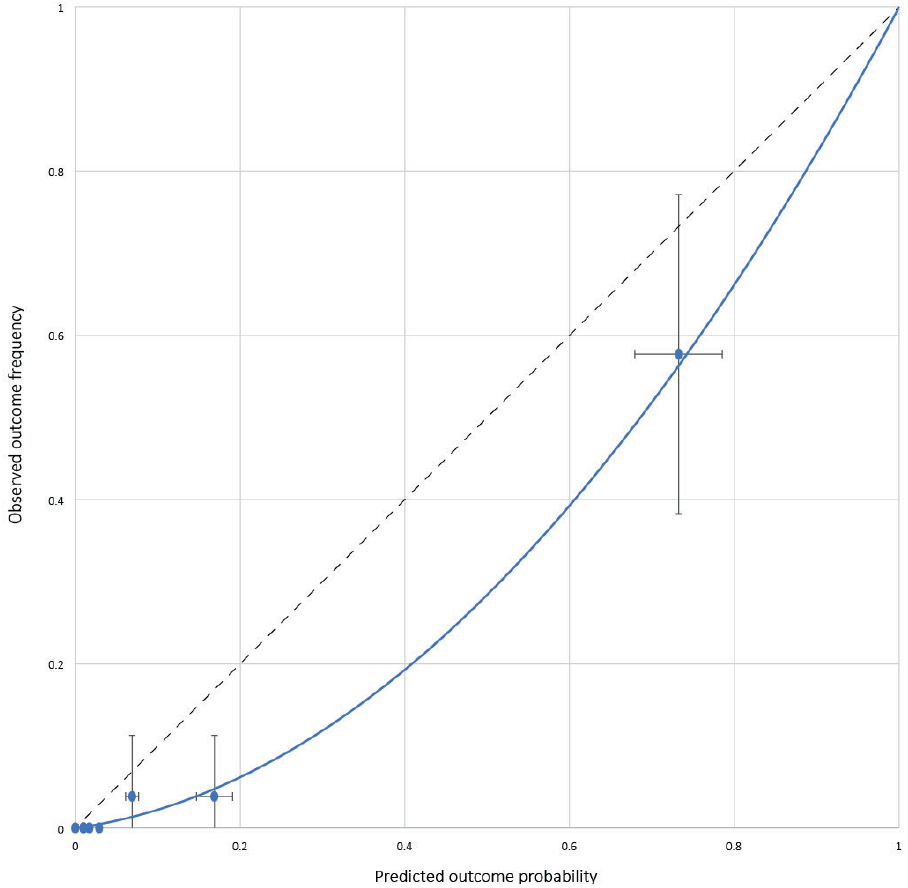
Calibration plot of the full model, analysed in a single-imputed dataset.

**Figure 3.**
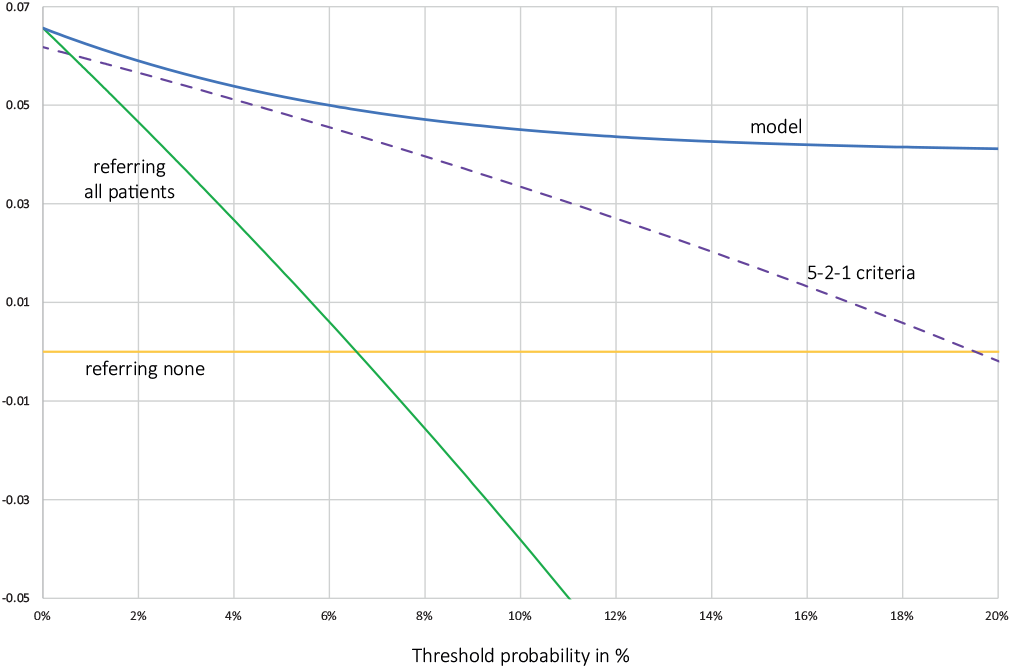
Decision curve analysis. The figure displays the net benefit for the full model and the 5-2-1criteria, while it also shows the net benefit for referring either all patients or none.

**Figure 4.**
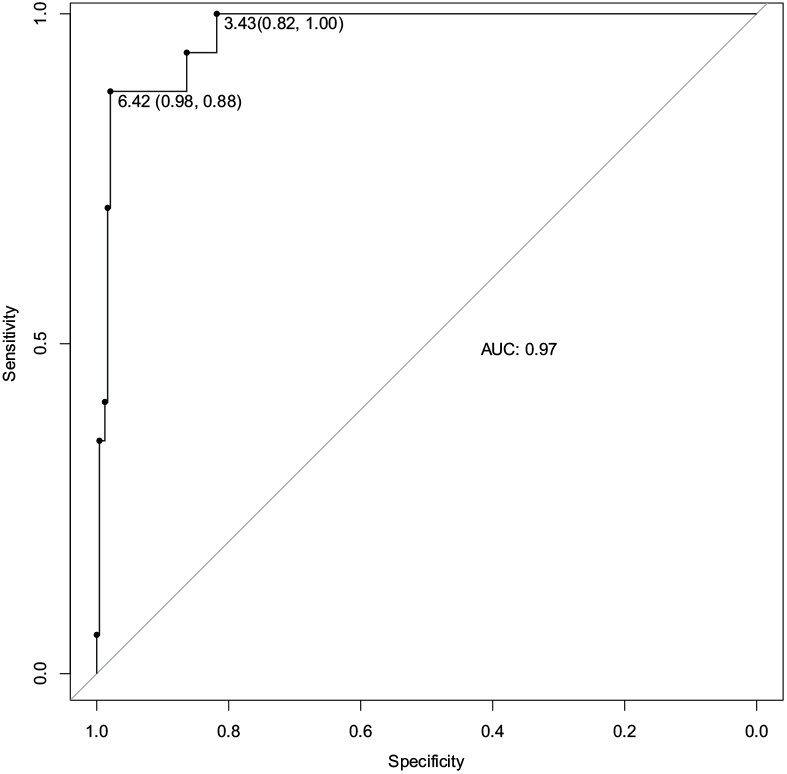
Receiver operating characteristics (ROC) curve of the screening tool (simplified scoring system).

Based on our arbitrary definition, the optimal cut-off point of the screening tool was 6.42. Using this cut-off point, the screening tool had a higher positive predictive value than the 5-2-1 criteria (76 vs. 20% respectively; table 5). Both models had a comparable sensitivity (88 vs 94%), while the screening tool had a higher specificity than the 5-2-1 criteria (98 vs 73%).

**Table 5.**
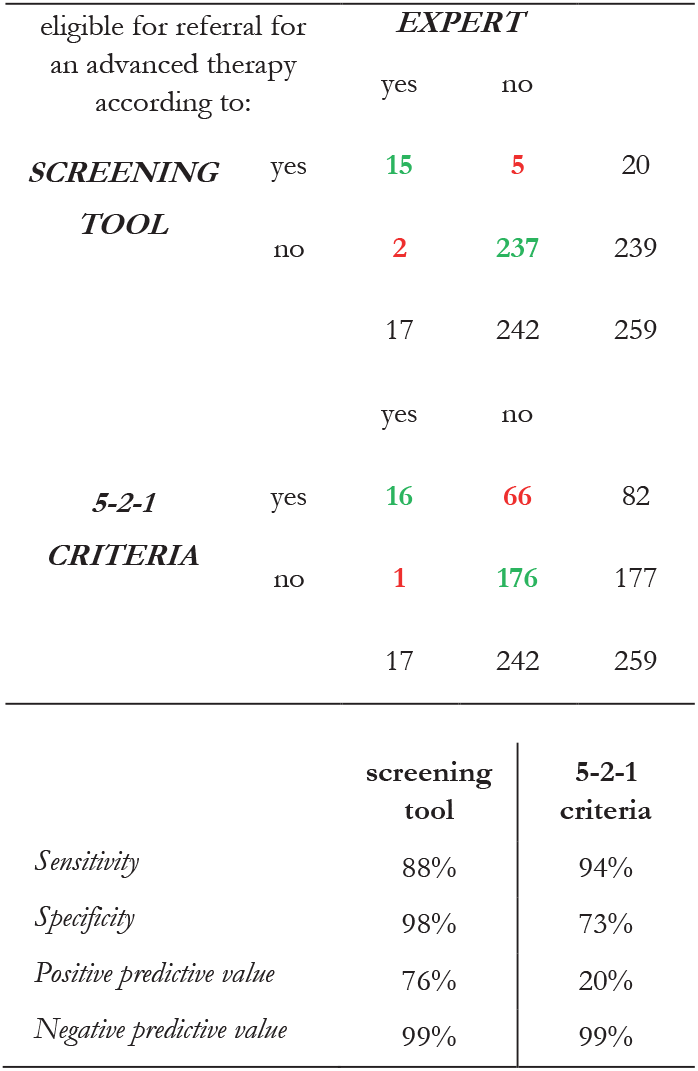
Eligibility for referral for an advanced therapy according to the screening tool and the 5-2-1 criteria. Expert refers to the reference test.

|  | eligible for referral for an advanced therapy according to: | EXPERT |  |  |
| --- | --- | --- | --- | --- |
|  |  | yes | no |  |
| SCREENING TOOL | yes | 15 | 5 | 20 |
|  | no | 2 | 237 | 239 |
|  |  | 17 | 242 | 259 |
| 5-2-1 CRITERIA | yes | 16 | 66 | 82 |
|  | no | 1 | 176 | 177 |
|  |  | 17 | 242 | 259 |

## DISCUSSION

This study had a number of important results. First, the prevalence of eligibility for referral for AT was relatively low, being 6.6% in a population of consecutive PD patients in a secondary care setting. Second, this study showed several strong predictors of eligibility for referral for AT, such as the presence of response fluctuations and troublesome dyskinesias. Finally, the results enabled the development of a three-factor screening tool (LEDD, response fluctuations, troublesome dyskinesias). This tool outperformed the 5-2-1 criteria.

The primary aim of this study was to develop a screening tool that helps in determining whether a PD patient should be referred to an specialized clinic for considering treatment with AT. This goal was not revolutionary, as the first attempts to improve the referral for AT go back to 2004, when the FLASQ-PD was designed to screen for potential DBS candidates.[27] Since then, several studies have been published to provide guidance to general neurologists to determine which PD patient might benefit from AT, either using clinical criteria or using wearable sensors.[5,13–15,28–30] However, most papers were focused on referral criteria for DBS only. In addition, referral criteria were based on expert consensus or assessment of fictitious cases by a group of experts.[10,11] The present study, however, used a different approach, as it investigated eligibility for referral for at least one AT by studying an unselected group of consecutive PD patients in routine care. An advantage of this methodology was the possibility to estimate the prevalence of referral eligibility, and thus to obtain a reliable estimation of the positive predictive value.

During the data collection of this study, Antonini *et al*. presented a consensus statement of international experts that eventually led to the development of the so-called 5-2-1 criteria.[14,31] The 5-2-1 criteria were introduced to help general neurologists identify advanced PD patients who are possibly eligible for AT.[17] These criteria are easy to use and the name itself serves as a mnemonic device. Our study data showed that the 5-2-1 criteria have an almost perfect sensitivity, but a considerable number of false positives.

In the present study, we developed a new model using empirical data and adequate statistical techniques. The variables in our final model partly corroborate the items included in the 5-2-1 criteria. Both the 5-2-1 criteria and our model include presence of dyskinesias and a measure of medication use (frequency of drugs vs. LEDD, respectively). However, the 5-2-1 includes «at least 2 hours *off* time per day» while this variable was excluded from our model during statistical item selection. Instead, our model includes presence of response fluctuations, which implicitly refers to presence of *off* periods.

Despite the similarities between the two models, decision curve analysis (DCA) showed that our screening tool had a higher net benefit than the 5-2-1 criteria for all possible threshold values. Nevertheless, both the screening tool and the 5-2-1 criteria had similar testing characteristics for low threshold values of the screening tool. The threshold value of DCA is informative of how one weighs the relative ‘costs’ of false-positive and false-negative results (e.g. a threshold of 5% is similar to saying “not referring a patient (false negative) is 19 times worse than referring a patient unnecessarily”). [24,26] Thus, both models are comparable when the costs of false negatives are considerably lower than the costs of inappropriate referrals. But in the absence of empirical data on the clinical utility of referring patients for AT, it remains unknown how one should weigh such costs. Therefore, it is not yet possible to decide on the optimal threshold value for referring patients.

For the purpose of illustration, we arbitrarily chose a cut-off point for our screening tool by maximising the sum of sensitivity and specificity. This resulted in a positive predictive value of 76% but an estimated sensitivity of 88%, being slightly lower that the 5-2-1 criteria. However, using the model is an iterative process. Importantly, during a subsequent check-up with slight progression of disease the patients initially ‘overlooked’ using the screening tool may well have become eligible and still being in time for subsequent AT.

This study had some limitations. First, the reference test, for which we used an expert consensus, was suboptimal. The experts had to judge eligibility for referral based on the information from the study assessments forms, as filled out by the general neurologist. Possibly, the experts would judge differently if they could have examined the patients themselves. However, as the outcome of this study was eligibility for referral, a fair judgement is based on referral information only. Another limitation is that the model has not yet been validated in a different population. Currently, our research team is about to conduct a validation study in another region in the Netherlands. Validation of the screening tool is essential prior to application in daily practice.

Future studies do not only need to validate the newly-developed screening tool, but also need to determine whether patients will actually benefit from the screening tool. In addition, future studies should include patient’s expectations about the treatment options, as it was recently shown that patient’s expectation are a good predictor of eligibility for DBS.[32] Moreover, new insights on the timing of AT, for example the expected results from the EARLY-PUMP trial studying earlier application of CSAI,[33] might require revision of the developed screening tool and other selection criteria.

In conclusion, we developed a prototype screening tool to screen for PD patients being eligible for referral for AT. Our tool appeared to outperform the 5-2-1 criteria. We expect that, after external validation of the model, implementation of the new screening tool will contribute to better and more timely recognition of patients who are eligible for referral for AT. This will lead to a more efficient referral policy, as more patients will be referred who were otherwise not recognized, while the number of inappropriate referrals will decrease.

## Data Availability

Data are available upon reasonable request.

## SUPPLEMENTARY INFORMATION

Tables and figures referenced in the main text are listed in the appendix. The TRIPOD checklist is also included in the appendix.

## ACKNOWLEDGMENTS

The authors thank all patients, general neurologists and Parkinson nurses who participated in this study.

In addition, the authors thank Wim Simons and Martin van Schijndel from the Dutch Parkinson Patient ‘s Association for their critical reflections on this design and results of this study and for their feedback on an earlier version of this manuscript.

Thanks to Paulus Bax for his valuable comments on the statistical analyses. We thank Claudia B. Gremmer for digitalizing and abstracting the study documents.

---

**Table A1.**
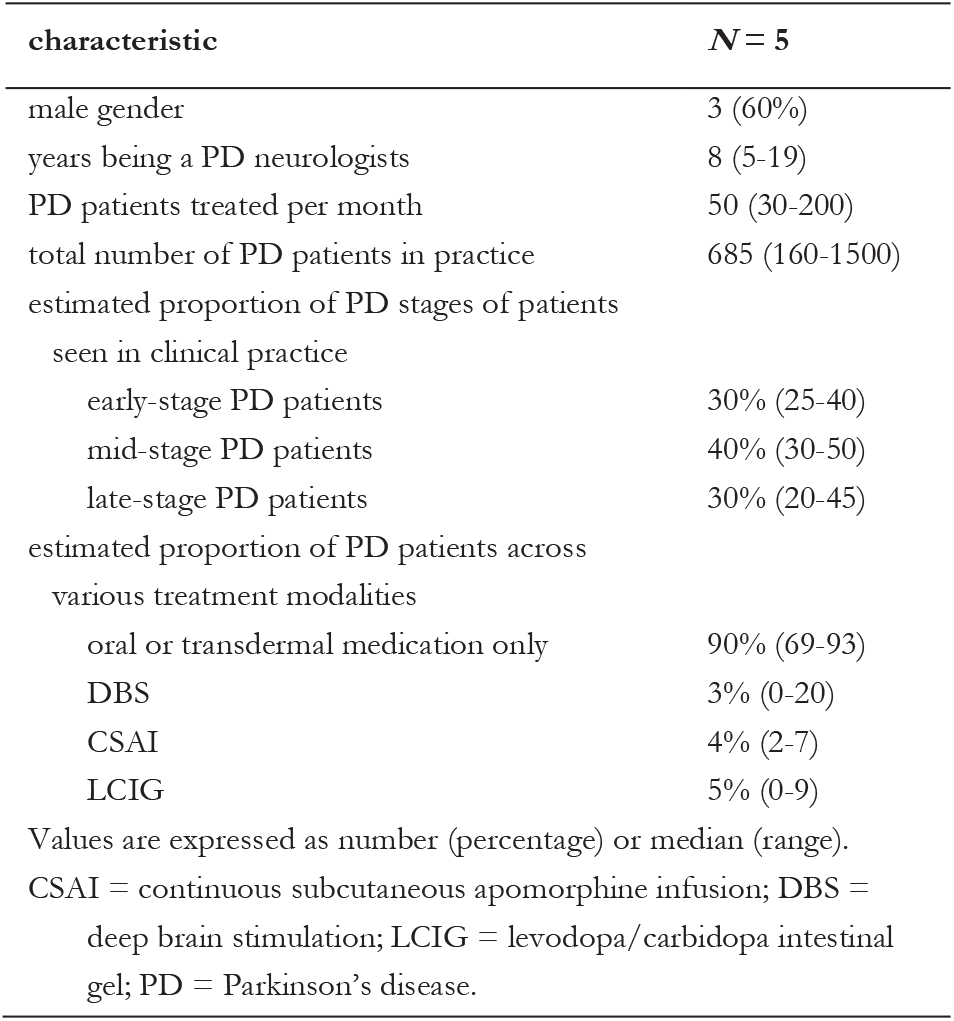
Characteristics of the members of the expert panel

**Figure A1.**
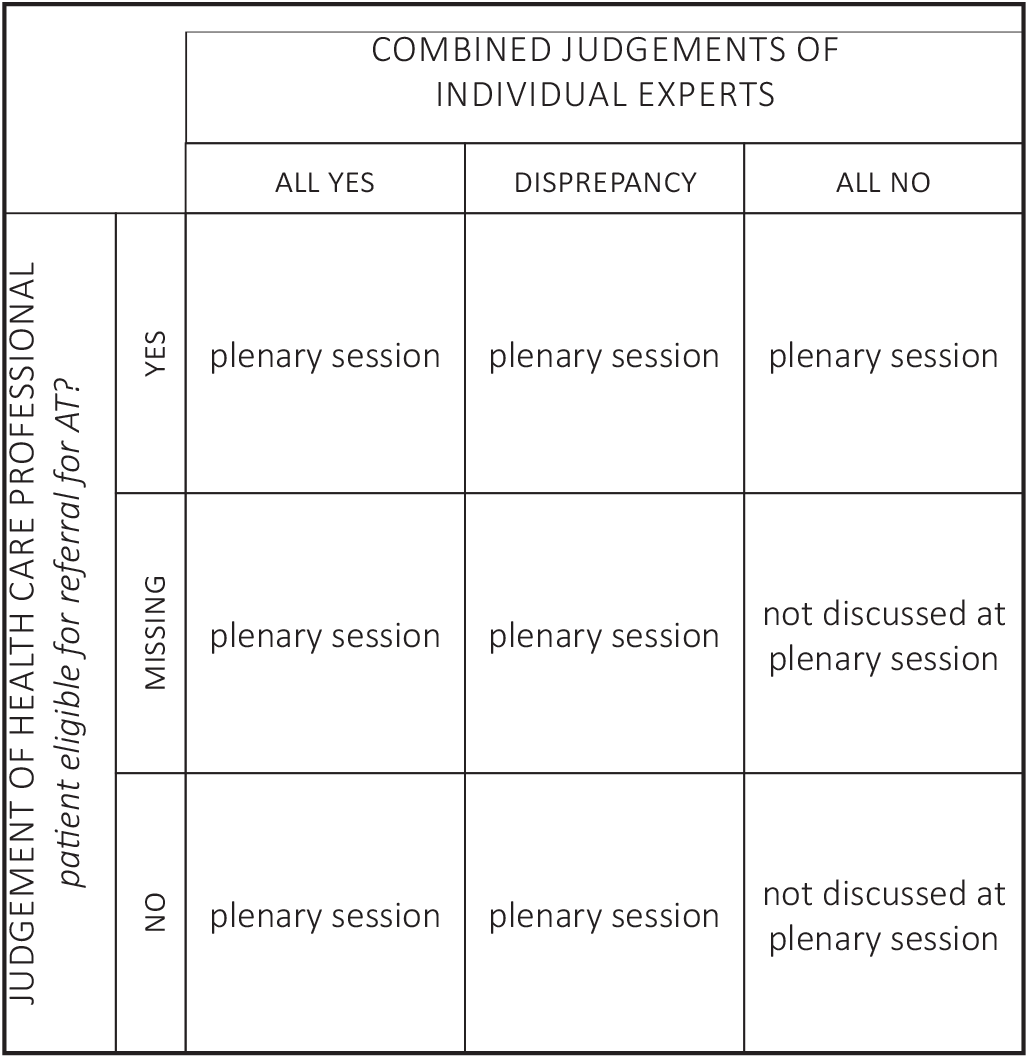
Cases selected for a plenary session with members of the expert panel

**Table A2.** All potential predictors for eligibility for referral for an advanced therapy

| predictor | source | comment |
| --- | --- | --- |
| sex | AF | male; female |
| age (years) | AF; f | [date] – [birth date] \| birth date was derived from the year of birth. In all patients, the day of birth was arbitrarily set on July 1 from the respective year of birth. Please note: to act according to current privacy legislation, we only included year of birth on the assessment form, not the exact birth date. |
| duration of disease (years) | AF; f | [date] – [date of diagnosis] \| the date of diagnosis was derived from the year of diagnosis. In all patients, the date of diagnosis was arbitrarily set on July 1 in the year in which the diagnosis PD was made. |
| total score NMS (points) | NMS | calculated in patients who filled out all items; in cases items were missing, the total score was not calculated. |
| LEDD (mg/day) | AF; f | calculated with the formula as described by Esselink *et al.* [23] |
| oral dopaminergic drug doses (n/day) | AF; f | times a day that the patient takes oral dopaminergic medication; this number did only include the planned number of medication moments, not any optional (symptom-dependent) intakes. |
| partner/relationship | AF | yes; no |
| lives independently | AF | yes; no |
| comorbidity | AF | no comorbidity; at least one comorbidity |
| response fluctuations | AF | none; predictable; unpredictable. |
| therapy resistant tremor | AF | yes; no |
| troublesome dyskinesias (hours/day) | AF | none; 0-2 hours/day; 2-4 hours/day; >4 hours/day |
| off periods (hours/day) | AF | none; 0-2 hours/day; 2-4 hours/day; >4 hours/day. |
| level of ADL impairment | AF | none; mild; moderate; severe. |
| visual hallucinations | AF | yes; no |
| daytime sleepiness | AF | yes; no |
| orthostatic hypotension | AF | yes; no |
| impulse control disorders | AF | yes; no |
| depression/anxiety | AF | yes; no |
| dysarthria during on | AF | yes; no |
| freezing during on | AF | yes; no |

**Table A2** All potential predictors for eligibility for referral for an advanced therapy (*continued*)
| predictor | source | comment |
| --- | --- | --- |
| PD dementia | AF | none; mild; moderate; severe |
| dribbling of saliva during daytime | NMS | yes; no [question 1] |
| loss or change in taste or smell | NMS | yes; no [question 2] |
| dysphagia | NMS | yes; no [question 3] |
| vomiting or nausea | NMS | yes; no [question 4] |
| constipation | NMS | yes; no [question 5] |
| faecal incontinence | NMS | yes; no [question 6] |
| incomplete bowel emptying | NMS | yes; no [question 7] |
| urge incontinence for urine | NMS | yes; no [question 8] |
| nocturia | NMS | yes; no [question 9] |
| unexplained pains | NMS | yes; no [question 10] |
| unexplained change in weight | NMS | yes; no [question 11] |
| problems remembering things | NMS | yes; no [question 12] |
| anhedonia | NMS | yes; no [question 13] |
| visual or auditory hallucinations | NMS | yes; no [question 14] |
| difficulty concentrating | NMS | yes; no [question 15] |
| feeling sad or 'low' | NMS | yes; no [question 16] |
| anxiety | NMS | yes; no [question 17] |
| feeling less or more interested in sex | NMS | yes; no [question 18] |
| difficulty to have sex | NMS | yes; no [question 19] |
| signs of orthostatic hypotension | NMS | yes; no [question 20] |
| falling | NMS | yes; no [question 21] |
| difficulty to stay awake during activities | NMS | yes; no [question 22] |
| difficulty getting to sleep at night | NMS | yes; no [question 23] |
| intense, vivid dreams | NMS | yes; no [question 24] |
| talking or moving during sleep (sign of RBD) | NMS | yes; no [question 25] |
| restless legs | NMS | yes; no [question 26] |
| swelling of legs | NMS | yes; no [question 27] |
| excessive sweating | NMS | yes; no [question 28] |
| double vision | NMS | yes; no [question 29] |
| delusions | NMS | yes; no [question 30] |
| AF = assessment form (filled out by general neurologist or Parkinson nurse); f = formula/calculation; NMS = NMS questionnaire (filled out by patient); LEDD = levodopa equivalent daily dose; PD = Parkinson's disease; RBD = REM-sleep behaviour disorder |  |  |

**Figure A2.**
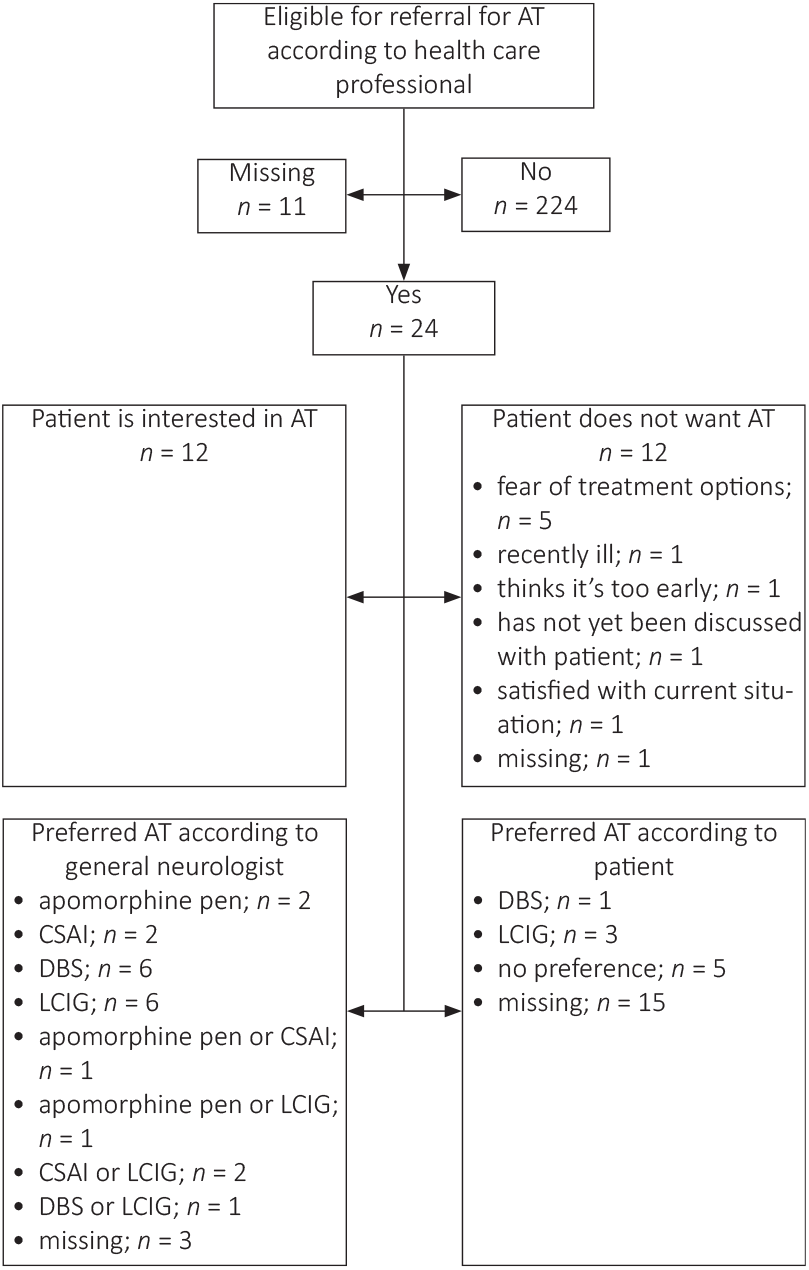
Figure displays eligibility according to health care professional (general neurologist or supervised Parkinson nurse), whether the patient is interested an advanced therapy, and preferred advanced therapy. AT = advanced therapy; CSAI = continuous subcutaneous apomorphine infusion; DBS = deep brain stimulation; LCIG = levodopa/carbidopa intestinal gel.

**Figure A3.**
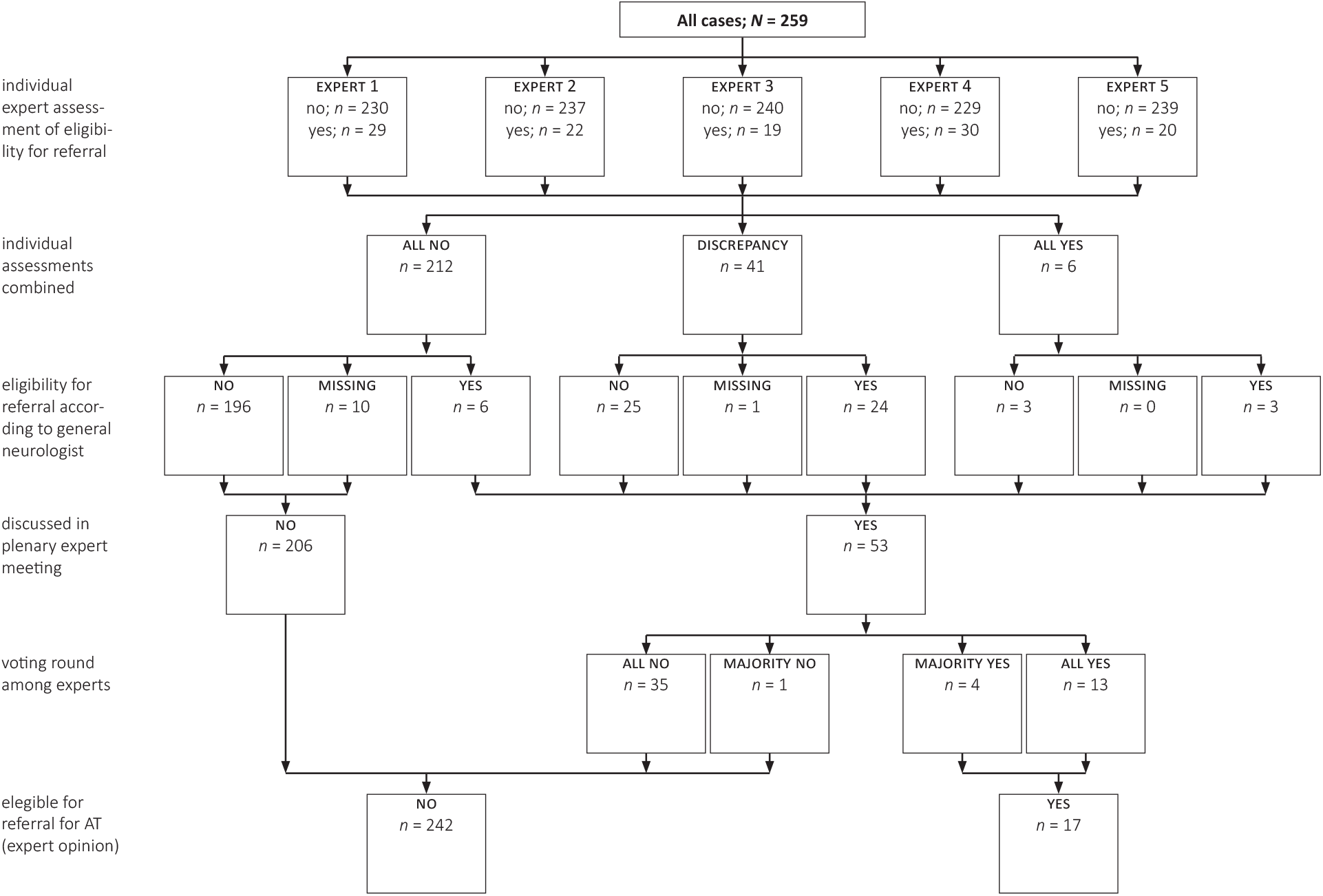
A staged approach was used for assessment of eligibility for referral. First, all experts assessed all cases individually. Cases that were deemed ineligible by all experts *and* by the general neurologists, were not discussed in the plenary expert meeting. All other cases were discussed in a plenary meeting. AT = advanced therapy.

**Table A3.** Eligibility for referral for an advanced therapy according to reference test (expert consensus) compared to assessment by the referring health care professional

|  | eligible for referral for AT according to: | *EXPERT* |  |  |
| --- | --- | --- | --- | --- |
|  |  | yes | no |  |
| *HEALTH CARE PROFESSIONAL\*\** | yes | 10 | 14 | 24 |
|  | no | 7 | 137 | 224 |
|  |  | 17 | 231 | 248* |

\* N = 248 due to 11 missing assessments by the health care professional.
\*\* General neurologist or Parkinson nurse.
Cohen's kappa: 0.44

**Table A4.** Scores on items of the NMS questionnaire of the included cases

| characteristic | all cases<br>(*N* = 259) | eligible for<br>referral for AT<br>(*N* = 17) | not eligible for<br>referral for AT<br>(*N* = 242) | missing, *n* |
| --- | --- | --- | --- | --- |
| NMS quest total score | 7.9 (SD: 4.6; [0-25]) | 9.2 (SD: 4.9) | 7.8 (SD: 4.5) | 52 |
| 1. dribbling of saliva during daytime | 101 (39.1%) | 3 (17.6%) | 98 (40.7%) | 1 |
| 2. loss or change in taste or smell | 70 (27.2%) | 6 (35.3%) | 64 (26.7%) | 2 |
| 3. dysphagia | 64 (24.7%) | 7 (41.2%) | 57 (23.6%) | 0 |
| 4. vomiting or nausea | 22 (8.6%) | 3 (17.6%) | 19 (7.9%) | 2 |
| 5. constipation | 88 (34.1%) | 7 (41.2%) | 81 (33.6%) | 1 |
| 6. faecal incontinence | 23 (9.0%) | 3 (17.6%) | 20 (8.4%) | 3 |
| 7. incomplete bowel emptying | 42 (16.4%) | 1 (5.9%) | 41 (17.2%) | 3 |
| 8. urge incontinence for urine | 138 (54.1%) | 10 (58.8%) | 128 (53.8%) | 4 |
| 9. nocturia | 136 (52.7%) | 11 (64.7%) | 125 (51.9%) | 1 |
| 10. unexplained pains | 55 (21.4%) | 5 (31.3%) | 50 (20.7%) | 2 |
| 11. unexplained change in weight | 39 (15.2%) | 7 (41.2%) | 32 (13.4%) | 3 |
| 12. problems remembering things | 108 (42.2%) | 8 (47.1%) | 100 (41.8%) | 3 |
| 13. anhedonia | 66 (25.5%) | 3 (17.6%) | 63 (26.0%) | 0 |
| 14. visual or auditory hallucinations | 51 (19.8%) | 5 (29.4%) | 46 (19.2%) | 2 |
| 15. difficulty concentrating | 131 (50.6%) | 7 (41.2%) | 124 (51.2%) | 0 |
| 16. feeling sad or 'low' | 78 (30.2%) | 5 (29.4%) | 73 (30.3%) | 1 |
| 17. anxiety | 40 (15.5%) | 3 (17.6%) | 37 (15.4%) | 1 |
| 18. feeling less interested or more interested in sex | 55 (22.3%) | 1 (7.1%) | 54 (23.2%) | 12 |
| 19. difficulty to have sex | 66 (28.3%) | 2 (14.3%) | 64 (29.2%) | 26 |
| 20. signs of orthostatic hypotension | 119 (46.1%) | 10 (58.8%) | 109 (45.2%) | 1 |
| 21. falling | 56 (21.6%) | 7 (41.2%) | 49 (20.2%) | 0 |
| 22. difficulty to stay awake during activities | 41 (15.8%) | 1 (5.9%) | 40 (16.5%) | 0 |
| 23. difficulty getting to sleep at night | 81 (31.8%) | 5 (29.4%) | 76 (31.9%) | 4 |
| 24. intense, vivid dreams | 77 (30.0%) | 4 (25.0%) | 73 (30.3%) | 2 |
| 25. signs of RBD | 96 (38.4%) | 7 (43.8%) | 89 (38.0%) | 9 |
| 26. restless legs | 102 (39.7%) | 10 (58.8%) | 92 (38.3%) | 2 |
| 27. swelling of legs | 49 (18.9%) | 3 (17.6%) | 46 (19.0%) | 0 |
| 28. excessive sweating | 48 (18.5%) | 7 (41.2%) | 41 (16.9%) | 0 |
| 29. double vision | 27 (10.5%) | 3 (17.6%) | 24 (10.0%) | 3 |
| 30. delusions | 20 (7.8%) | 1 (5.9%) | 19 (7.9%) | 2 |
| Values expressed as number (percentage of complete cases) or mean (SD; [lowest-highest]). |  |  |  |  |
| AT = advanced therapy; NMS = non-motor symptoms; RBD = REM-sleep behaviour disorder. |  |  |  |  |

**Table A5.** Univariable logistic regression analysis of the NMS questionnaire

| characteristic | odds ratio<br>(95% CI) | *p*-value* | complete/missing |  |
| --- | --- | --- | --- | --- |
|  |  |  | eligible<br>(*N* = 17) | not eligible<br>(*N* = 242) |
| NMS quest total score | 1.06 (0.95-1.20) | 0.310 | 12/5 | 195/47 |
| 1. dribbling of saliva during daytime | 0.31 (0.08-1.12) | 0.074 | 17/0 | 241/1 |
| 2. loss or change in taste or smell | 1.50 (0.53-4.22) | 0.443 | 17/0 | 240/2 |
| 3. dysphagia | 2.27 (0.83-6.24) | 0.111 | 17/0 | 242/0 |
| 4. vomiting or nausea | 2.49 (0.66-9.44) | 0.179 | 17/0 | 240/2 |
| 5. constipation | 1.38 (0.51-3.77) | 0.526 | 17/0 | 241/1 |
| 6. faecal incontinence | 2.35 (0.62-8.86) | 0.208 | 17/0 | 239/3 |
| 7. incomplete bowel emptying | 0.30 (0.04-2.34) | 0.252 | 17/0 | 239/3 |
| 8. urge incontinence for urine | 1.23 (0.45-3.33) | 0.687 | 17/0 | 238/4 |
| 9. nocturia | 1.70 (0.61-4.75) | 0.310 | 17/0 | 241/1 |
| 10. unexplained pains | 1.74 (0.58-5.23) | 0.326 | 16/1 | 241/1 |
| 11. unexplained change in weight | 4.53 (1.61-12.75) | **0.004** | 17/0 | 239/3 |
| 12. problems remembering things | 1.24 (0.46-3.31) | 0.674 | 17/0 | 239/3 |
| 13. anhedonia | 0.61 (0.17-2.19) | 0.447 | 17/0 | 242/0 |
| 14. visual or auditory hallucinations | 1.76 (0.59-5.24) | 0.311 | 17/0 | 240/2 |
| 15. difficulty concentrating | 0.67 (0.25-1.81) | 0.425 | 17/0 | 242/0 |
| 16. feeling sad or 'low' | 0.96 (0.33-2.82) | 0.939 | 17/0 | 241/1 |
| 17. anxiety | 1.18 (0.32-4.31) | 0.801 | 17/0 | 241/1 |
| 18. feeling less or more interested in sex | 0.26 (0.03-1.99) | 0.193 | 14/3 | 233/9 |
| 19. difficulty to have sex | 0.40 (0.09-1.86) | 0.244 | 14/3 | 219/23 |
| 20. signs of orthostatic hypotension | 1.73 (0.64-4.70) | 0.282 | 17/0 | 241/1 |
| 21. falling | 2.76 (1.00-7.61) | **0.050** | 17/0 | 242/0 |
| 22. difficulty to stay awake during activities | 0.32 (0.04-2.45) | 0.270 | 17/0 | 242/0 |
| 23. difficulty getting to sleep at night | 0.89 (0.30-2.61) | 0.829 | 17/0 | 238/4 |
| 24. intense, vivid dreams | 0.77 (0.24-2.46) | 0.655 | 16/1 | 241/1 |
| 25. signs of RBD | 1.27 (0.46-3.52) | 0.650 | 16/1 | 234/8 |
| 26. restless legs | 2.30 (0.85-6.25) | 0.103 | 17/0 | 240/2 |
| 27. swelling of legs | 0.91 (0.25-3.31) | 0.890 | 17/0 | 242/0 |
| 28. excessive sweating | 3.43 (1.23-9.54) | **0.018** | 17/0 | 242/0 |
| 29. double vision | 1.92 (0.52-7.16) | 0.332 | 17/0 | 239/3 |
| 30. delusions | 0.73 (0.09-5.78) | 0.763 | 17/0 | 240/2 |

\* *P*-values smaller than *p* < 0.05 are marked in red.
NMS = non-motor symptoms; RBD = REM-sleep behaviour disorder.

TRIPOD Checklist: Prediction Model Development and Validation

| Section/Topic | Item |  | Checklist Item | Page |
| --- | --- | --- | --- | --- |
| Title and abstract |  |  |  |  |
| Title | 1 | D;V | Identify the study as developing and/or validating a multivariable prediction model, the target population, and the outcome to be predicted. | 1 |
| Abstract | 2 | D;V | Provide a summary of objectives, study design, setting, participants, sample size, predictors, outcome, statistical analysis, results, and conclusions. | 1 |
| Introduction |  |  |  |  |
| Background and objectives | 3a | D;V | Explain the medical context (including whether diagnostic or prognostic) and rationale for developing or validating the multivariable prediction model, including references to existing models. | 2 |
|  | 3b | D;V | Specify the objectives, including whether the study describes the development or validation of the model or both. | 2 |
| Methods |  |  |  |  |
| Source of data | 4a | D;V | Describe the study design or source of data (e.g., randomized trial, cohort, or registry data), separately for the development and validation data sets, if applicable. | 2 |
|  | 4b | D;V | Specify the key study dates, including start of accrual; end of accrual; and, if applicable, end of follow-up. | 2 |
| Participants | 5a | D;V | Specify key elements of the study setting (e.g., primary care, secondary care, general population) including number and location of centres. | 2 |
|  | 5b | D;V | Describe eligibility criteria for participants. | 3 |
|  | 5c | D;V | Give details of treatments received, if relevant. | n.a. |
| Outcome | 6a | D;V | Clearly define the outcome that is predicted by the prediction model, including how and when assessed. | 3 |
|  | 6b | D;V | Report any actions to blind assessment of the outcome to be predicted. | 3 |
| Predictors | 7a | D;V | Clearly define all predictors used in developing or validating the multivariable prediction model, including how and when they were measured. | 3 |
|  | 7b | D;V | Report any actions to blind assessment of predictors for the outcome and other predictors. | 3 |
| Sample size | 8 | D;V | Explain how the study size was arrived at. | 4 |
| Missing data | 9 | D;V | Describe how missing data were handled (e.g., complete-case analysis, single imputation, multiple imputation) with details of any imputation method. | 4 |
| Statistical analysis methods | 10a | D | Describe how predictors were handled in the analyses. | 4 |
|  | 10b | D | Specify type of model, all model-building procedures (including any predictor selection), and method for internal validation. | 4 |
|  | 10c | V | For validation, describe how the predictions were calculated. | n.a. |
|  | 10d | D;V | Specify all measures used to assess model performance and, if relevant, to compare multiple models. | 4,5 |
|  | 10e | V | Describe any model updating (e.g., recalibration) arising from the validation, if done. | n.a. |
| Risk groups | 11 | D;V | Provide details on how risk groups were created, if done. | - |
| Development vs. validation | 12 | V | For validation, identify any differences from the development data in setting, eligibility criteria, outcome, and predictors. | n.a. |
| Results |  |  |  |  |
| Participants | 13a | D;V | Describe the flow of participants through the study, including the number of participants with and without the outcome and, if applicable, a summary of the follow-up time. A diagram may be helpful. | 5 |
|  | 13b | D;V | Describe the characteristics of the participants (basic demographics, clinical features, available predictors), including the number of participants with missing data for predictors and outcome. | 5 |
|  | 13c | V | For validation, show a comparison with the development data of the distribution of important variables (demographics, predictors and outcome). | nxa |
| Model development | 14a | D | Specify the number of participants and outcome events in each analysis. | 6 |
|  | 14b | D | If done, report the unadjusted association between each candidate predictor and outcome. | 6,7 |
| Model specification | 15a | D | Present the full prediction model to allow predictions for individuals (i.e., all regression coefficients, and model intercept or baseline survival at a given time point). | 8 |
|  | 15b | D | Explain how to the use the prediction model. | 8 |
| Model performance | 16 | D;V | Report performance measures (with CIs) for the prediction model. | 7,8 |
| Model-updating | 17 | V | If done, report the results from any model updating (i.e., model specification, model performance). | n.a. |
| Discussion |  |  |  |  |
| Limitations | 18 | D;V | Discuss any limitations of the study (such as nonrepresentative sample, few events per predictor, missing data). | 9,10 |
| Interpretation | 19a | V | For validation, discuss the results with reference to performance in the development data, and any other validation data. | n.a. |
|  | 19b | D;V | Give an overall interpretation of the results, considering objectives, limitations, results from similar studies, and other relevant evidence. | 9 |
| Implications | 20 | D;V | Discuss the potential clinical use of the model and implications for future research. | 10 |
| Other information |  |  |  |  |
| Supplementary information | 21 | D;V | Provide information about the availability of supplementary resources, such as study protocol, Web calculator, and data sets. | 10 |
| Funding | 22 | D;V | Give the source of funding and the role of the funders for the present study. | 10 |

\*Items relevant only to the development of a prediction model are denoted by D, items relating solely to a validation of a prediction model are denoted by V, and items relating to both are denoted D;V. We recommend using the TRIPOD Checklist in conjunction with the TRIPOD Explanation and Elaboration document.

